# Spatiotemporal dynamics of dengue in Colombia in relation to the combined effects of local climate and ENSO

**DOI:** 10.1101/2020.08.24.20181032

**Authors:** Estefanía Muñoz, Germán Poveda, M. Patricia Arbeláez, Iván D. Vélez

## Abstract

Dengue virus (DENV) is an endemic disease in the hot and humid low-lands of Colombia. We characterize diverse temporal and spatial patterns of monthly series of dengue incidence in diverse regions of Colombia during the period 2007-2017 at different spatial scales, and their association with indices of El Niño/Southern Oscillation (ENSO) at the tropical Pacific and local climatic variables. For estimation purposes, we use linear analysis tools including lagged cross-correlations (Pearson test), cross wavelet analysis (wavelet cross spectrum, and wavelet coherence), as well as a novel nonlinear causality method, PCMCI, that allows identifying common causal drivers and links among high dimensional simultaneous and time-lagged variables. Our results evidence the strong association of DENV cases in Colombia with ENSO indices and with local temperature and rainfall. El Niño (La Niña) phenomenon is related to an increase (decrease) of dengue cases nationally and in most regions and departments, with maximum correlations occurring at shorter time lags in the Pacific and Andes regions, closer to the Pacific Ocean. This association is mainly explained by the ENSO-driven increase in temperature and decrease in rainfall, especially in the Andes and Pacific regions. The influence of ENSO is not stationary (there is a reduction of DENV cases since 2005) and local climate variables vary in space and time, which prevents to extrapolate results from one site to another. The association between DENV and ENSO varies at national and regional scales when data are disaggregated by seasons, being stronger in DJF and weaker in SON. Specific regions (Pacific and Andes) control the overall relationship between dengue dynamics and ENSO at national scale, and the departments of Antioquia and Valle del Cauca determine those of the Andes and Pacific regions, respectively. Cross wavelet analysis indicates that the ENSO-DENV relation in Colombia exhibits a strong coherence in the 12 to 16-months frequency band, which implies the frequency locking between the annual cycle and the interannual (ENSO) timescales. Results of nonlinear causality metrics reveal the complex concomitant effects of ENSO and local climate variables, while offering new insights to develop early warning systems for DENV in Colombia.

## Introduction

Dengue virus (DENV) is considered the most important vector-borne viral disease, infecting around 50 million people per year [1] and with an estimation of 3.9 billion people at risk of contracting dengue in over 128 countries [2]. It is the arboviral disease provoking more human morbidity and mortality [3-7]. DENV incidence has shown an evident increase in the last decades [4,8,9], which has been associated with population growth, urbanization, and climate change, extending the geographical range where it is viable [7,10,11]. In spite of many efforts to curtail and prevent the incidence, DENV has continued to expand in endemic and new areas [6,12]. Public expenditures to cover treatments have increased accordingly [13], so that the cost of dengue in 18 countries in 2015 amounted to USD 3.3 billion expressed in terms of the Purchasing Power Parity (PPP) [14].

DENV is transmitted by Aedes mosquitoes, mainly by *Aedes aegypti* [15-17], an endophilic mosquito that lives in tropical and subtropical regions [18]. Given that there is a good correlation between the incidence rate of DENV and both adult and egg count [19,20], the increase of DENV cases could be related to an increase in mosquitoes population. The spread of *Ae. aegypti* is attributed to anthropogenic and climate conditions [3,21-26], mainly affecting the most vulnerable human populations of low-income urban centers [8,27]. In spite that weather and climate are considered to play an important role in the temporal and spatial distribution of DENV, the identification and relative importance of climatic variables can be controversial [10,11,28] and may differ among environments [29,30]. The relationship between climate variability and the incidence of dengue has been evaluated in diverse sites, such as Mexico, Puerto Rico, Costa Rica, Indonesia, Australia, and Colombia [31-38]. These studies have found that, similar to cholera and malaria, dengue incidence exhibits seasonal cycles and inter-annual variability [39], which are often linked to climatic factors [33,36,40].

Mosquitoes breed in warm and wet regions, which explains the high number of cases of infected people in tropical regions [15]. Temperature impacts both the entomological parameters related to the mosquitoes’ life cycle [41] and the incubation period of DENV in female mosquitoes [42], while precipitation determines the availability of breeding sites. On the other hand, diverse entomological factors of *Ae. aegypti* development and life cycle have been associated to weather and climate anomalies, in particular air temperature and precipitation, brought about by El Niño/Southern Oscillation (ENSO), the most important modulator of climate variability at interannual timescales worldwide [10,43].

Knowledge about the spatial distribution of dengue is essential to understand virus dynamics and to put in place early warning systems and prevention and control programs [8]. As Colombia has a wide range of climate conditions due to its equatorial location and the presence of the Andes, a study at different spatial scales can help to develop and improve regional and local public health interventions, and focus prevention programs and surveillance measures in DENV-prone areas [27]. Besides, further understanding about the linkages between climate variability and DENV is likely to anticipate the effects of climate change [43], and provide clues to the scenarios of future ENSO events [10]. Although dengue has been studied in Colombia [34,37,44-48], spatiotemporal analyses relating climate and dengue cases have been carried out at local level, and few regional studies have considered the combined effects of climate variables and ENSO [47]. Therefore, the main objective of this paper is to characterize the temporal and spatial distribution of dengue incidence and their association with climatic variables and ENSO at different time and spatial scales in Colombia. In particular we aim to confirm the wave-like traveling pattern of DENV incidence across the different departments of Colombia, as found by Acosta [47]. This will help us to advance our understanding of DENV seasonal endemicity and epidemic outbreaks. This paper also analyzes the relationship between the incidence of DENV and the occurrence of both phases of ENSO: El Niøo (warm phase) and La Nina (cold phase), through the possible climate variables that are affected by these macro-climatic phenomena.

The work is distributed as follows. The next section describes the databases and the methods, the third section reviews some important remarks of climate and dengue in Colombia and shows the linkages between local climate, ENSO and dengue at national, regional, departmental, and local levels, fourth section discusses the results obtained and confronts them with the results of other studies, and the last section summarizes the main conclusions found with the analyzes performed.

## Materials and methods

### Study site

This paper evaluates the spatiotemporal dynamics of dengue incidence and its relationship with climatic variables and ENSO in Colombia. The country is located in northwestern tropical South America (13.4°N-4.2°S, 66.8°W-81.7°W), and it is divided into 5 major natural regions, namely Caribbean, Andes, Pacific, Orinoco and Amazon, 32 departments and 1122 municipalities, and has a population of more than 50 million people, of which about 24 million live in dengue-prone areas [49].

### Databases

Dengue data corresponds to daily dengue cases reported from 2007 to 2017 by the Public Health Surveillance System (Sistema de Vigilancia en Salud Pública, SIVIGILA, https://www.ins.gov.co/Direcciones/Vigilancia/Paginas/SIVIGILA.aspx) of the National Institute of Health (Instituto Nacional de Salud, INS) of Colombia. Information on each dengue case includes the date and the municipality in which the patient had the medical consultation. Such information contains confirmed and potential cases. The database has information from 1058 municipalities.

Climatic variables were provided by the governmental Instituto Colombiano de Hidrología, Meteorología y Estudios Ambientales (IDEAM). This database includes daily maximum temperature *(T_max_)*, minimum temperature (*T_min_*), precipitation (*P*), wind velocity (*WV*), and relative humidity *(HR)*. We selected stations with records that match the dengue database (2007-2017). In total, we used information from 1595 stations of *P*, 305 stations of *T_min_*, and 295 stations of *T_max_*. As few data are available regarding wind velocity (50 stations) and humidity (14 stations), and almost none completely overlap with the dengue series, we only considered these variables for analyzes at national level. Finally, we use the Oceanic Niño Index (ONI, https://origin.cpc.ncep.noaa.gov/products/analysis_monitoring/ensostuff/ONI_v5.php) as a variable that represents the dynamics of El Niøo/Southern Oscillation (ENSO) phenomenon. This index is based on a three-month running temperature mean of anomalies in the Niøo 3.4 region (5°N-5°S, 120°-170°W). Furthermore, the ENSO Precipitation Index (ESPI; http://eagle1.umd.edu/GPCP_ICDR/espi.htm) is also used to quantify non-linear causalities between ENSO, local climate and dengue incidence at national and regional levels.

## Methods

Data regarding dengue cases and climatic variables are aggregated at monthly scale, avoiding the presence of zeros in rainfall data at daily scale that affect correlation estimates. These data are also standardized through subtraction of the long-term monthly mean and scaling by the long-term monthly standard deviation. Standardization filters out part of the stationary cycles, such as the annual cycle, allowing the visualization of phenomena at interannual timescales.

For estimation purposes, we use linear analysis tools including lagged cross-correlations (Pearson test), and wavelet analysis, and diverse time-delayed nonlinear causal discovery metrics. Cross-correlations (*ρ*) between dengue cases and ONI, dengue cases and climatic series, and ONI and climatic series are estimated over a range of lags between 0 and 12 months. Lagged cross-correlograms are estimated by aggregating the information at national, regional, departmental and municipal scales. Furthermore, the correlations between raw dengue cases (without standardization) and ONI are performed by splitting the data into quarters (DJF, MAM, JJA, SON). This kind of analysis quantifies the degree of linear association and the time delay between the time-series.

We also perform wavelet analysis to study the time-frequency behavior of monthly series of dengue and ONI, and their conjoint dynamics at the national scale. This technique has the advantage to process scale-dependent non-stationary time series, thus allowing the description of the variability of the spectral properties over time. The coupled dynamics between ONI and dengue cases was evaluated using the wavelet coherence and the cross-wavelet transform. These analyses allow identifying time intervals and period bands in which two time-series are related [50,51]. This technique has already been used to analyze the dynamics of dengue in different places [11,25,33]. We implemented the continuous wavelet transform (CWT) based on Torrence and Compo [50] and the Cross Wavelet Analysis (CWA) based on Maraun and Kurths [52], using the waipy toolkit developed in Python, which was implemented by Mabel Calim Costa and available at https://github.com/mabelcalim/waipy.

Furthermore, we use diverse time-lagged causal inference methods aimed at discovering and quantifying the causal interdependencies between time series of weather variables, ENSO indices and dengue incidence at national and regional scales. To this end, we employ the PCMCI method which allows to identify common drivers and links among high dimensional time-lagged variables, by combining a PC Markovian condition-selection step, named after its creators Peter and Clark [53,54] and the Momentary Conditional Independence (MCI) test. PCMCI has been applied recently to a large suite of biogeophysical phenomena [55-60]. The PCMCI method uses diverse statistical tests to infer non-linear causalities including linear partial correlations (ParCorr) and three types of nonlinear independence tests: GPDC, CMI, and PCMCIplus. GPDC is based on Gaussian process regressions and a distance correlation test on the residuals, suitable for a large class of nonlinear dependencies with additive noise. CMI is a nonparametric test based on a k-nearest neighbor estimator of conditional mutual information that can accommodate almost any type of dependency [58-60]. PCMCIplus can identify the full, lagged and contemporaneous causal graph (up to the Markov equivalence class for contemporaneous causality) under the standard assumptions of Causal Sufficiency, and the Markov condition (J. Runge, pers. comm.). For implementation purposes, we use the Tigramite 4.2 python package, which allows to reconstruct graphical models (conditional independence graphs) from discrete or continuously-valued time series based on the PCMCI method and create high-quality plots of the results [58]. The module is available at https://github.com/jakobrunge/tigramite/.

### Dengue and climate

Climate can impact dengue dynamics via changes in pathogens, hosts, and transmission [61]. Temperature and precipitation are variables widely associated with dengue infection [24,41], being the peak incidence during the first (second) half of the year in the southern (northern) hemisphere in association with elevated temperature and precipitation [62,63]. High temperatures accelerate the metabolic rate of a vector, increasing biting rates of female mosquitoes and their longevity [28,64,65]. This may result in increases in population size and enhanced egg production. Temperature also influences the geographical range of vector survival [66] and humidity state. High humidity enhances adult mosquito survival, but although low humidity decreases the survival rate of arthropod vectors because of dehydration, and it also may cause an increase in the mosquito blood-feeding rate [67]. Precipitation provides breeding places [13,68], but the impacts depend on its interaction with evaporation [67], soil type, topography and the proximity of water bodies. Besides, heavy or prolonged rainfall events may disrupt vector breeding sites, and indeed, kill the mosquitoes directly [66].

Both the seasonality and interannual variability of dengue cases have shown connections with climate, existing considerable evidence of the role of El Niøo-Southern Oscillation (ENSO) on endemic infections [39,69,70]. ENSO is a macroclimatic phenomenon resulting from strong non-linear ocean-atmosphere feedbacks over the tropical Pacific, with an average frequency of 3-5 years, and constitutes the main modulator of climate variability at interannual timescales worldwide, with particular strong impacts in northern South America [71-73]. ENSO has two extremes phases known as El Niño (warm phase) and La Nina (cold phase), which take a huge toll in terms of human lives, socio-economic costs and environmental impacts worldwide. El Niño (La Niña) occurs with an average frequency of 3-4 (6-8) years and is associated with warmer (colder) sea surface temperature in the eastern and central equatorial Pacific Ocean [73].

Relationships between climate variables and factors affecting DENV transmission are complex, non-linear, and non-stationary [11,20,31,33,67]. Climate variables may impact the mosquito’s populations in different ways depending on local conditions since the fundamental processes of heat and water exchange are determined by the microclimate states [74]. Furthermore, ENSO’s impacts vary markedly, affected by the ENSO diversity and the modes of variability within and outside the tropical Pacific [73].

### Climate of Colombia

Colombia’s climate varies remarkably in time and space owing to its equatorial location and the temperature and rainfall gradients associated with the Andes topography. The temporal distribution of rainfall highly depends on the meridional oscillation of the Inter-Tropical Convergence Zone (ITCZ), the dynamics of three low-level jets: Choco, Caribbean and Orinoco [75-78], changes in topography (0 to 6,000 m), the ocean-atmosphere dynamics of the Pacific, Atlantic and Caribbean Sea, the Amazon and Orinoco River basins, and strong land surface-atmosphere feedbacks [79,80]. Mean annual temperature and rainfall depends on altitude, and geographic location. Mean annual rainfall ranges from 50 mm in deserts to 10,000-13,000 mm over the Pacific coast in one of the rainiest spots on Earth [81]. Besides, uni-modal and bi-modal annual cycles of precipitation vary throughout the different regions [22]. A bi-modal annual cycle is witnessed over the Andes of Colombia with wetter seasons in April-May and September-November and drier seasons during December-March and June-August. A uni-modal annual cycle predominates in other regions, with maximum and minimum rainfall rates depending on position of the ITCZ, and the aforementioned low-level jets and land-surface feedbacks. On the other hand, Colombia exhibits a strong interannual climatic variability associated with ENSO, showing prolonged dry (wet) periods and above (below) normal temperatures during El Niño (La Niña) [76,80,82-89].

Fig 1 shows the maximum cross-correlations *(ρ_max_)* between ONI and precipitation, ONI and maximum temperature, and ONI and minimum temperature and the associated lags. In general, the observed maximum correlation between ONI and precipitation in Colombia is negative *(ρ_max_* ≈-0.6, p ≤0.05), given that negative anomalies of precipitation (drier periods) occur during El Niño (higher ONI values).

**Fig 1.**
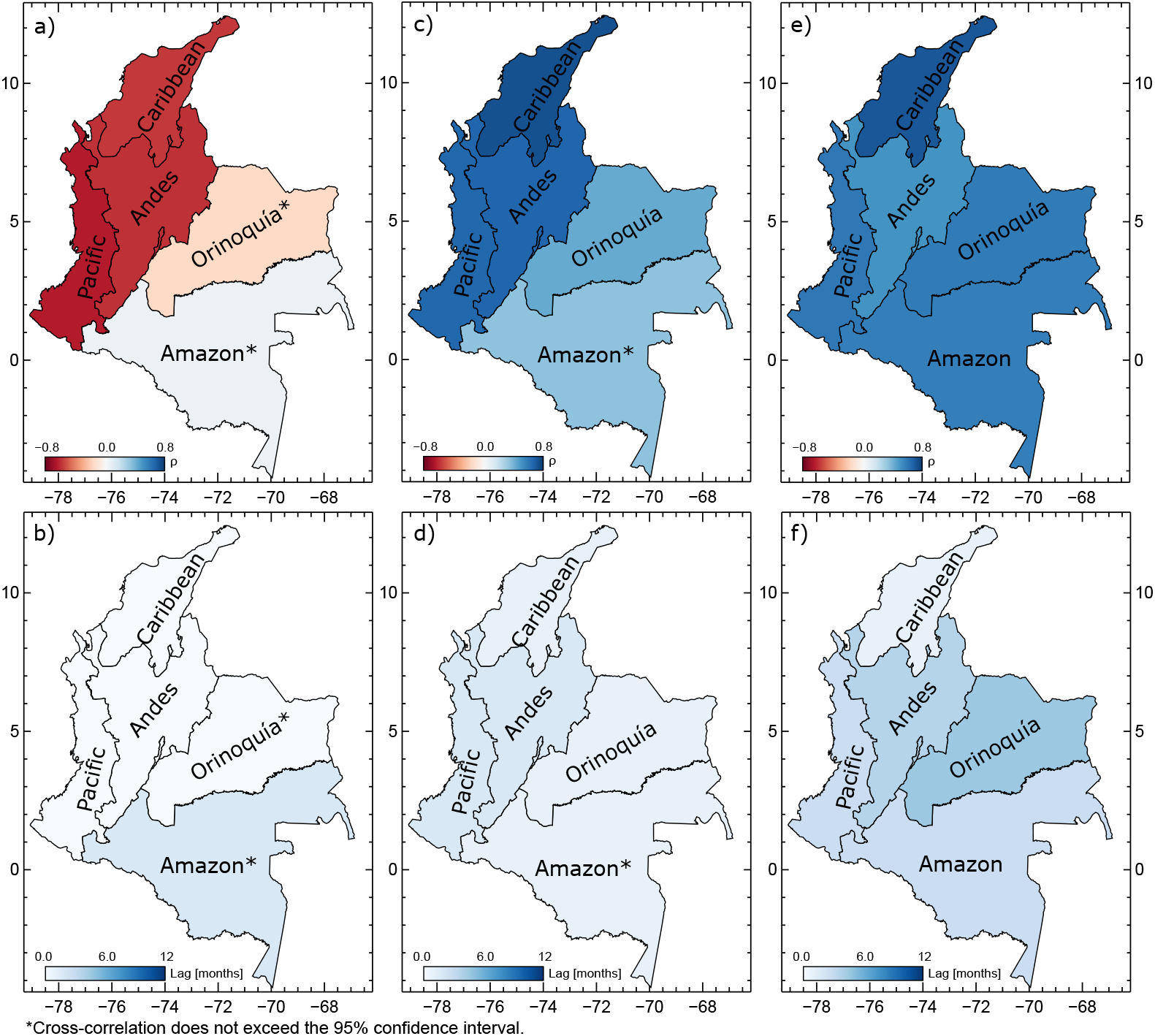
Cross-correlations between ONI and climate variables on the regional scale. Maximum cross-correlation *(p* ≤0.05) and lag between ONI and precipitation (a,b), maximum temperature (c,d), and minimum temperature (e,f).

Also, correlations between ONI and temperatures are positive *(ρ_max_* ≈ 0.75 for *T_max_* and *ρ_max_* ≈ 0.65 for *T_min_)*, denoting the increase in both temperatures during El Niøo. But there is variability among regions. Maximum linear correlations between ONI and P are simultaneous (lag 0) in all regions, except in the Amazon, which occurs at 2-months lag. Between ONI and *T_max_*, maximum correlation occur at 1-month lag for the Amazon, Orinoco, and Caribbean regions, and at 2-months lag for the Andes and Pacific regions, and between ONI and *T_min_*, at 1-month lag for the Caribbean, 3-months for the Pacific and Amazon, 4-months for the Andes, and 5-months for the Orinoco region.

The effects of ENSO are more noticeable in the Andes, Pacific, and Caribbean regions (see Fig 1a and Fig 1c). This is due to: (1) the weakening of the westerly low-level Chocó Jet [72,75,81,85,90,91]; (2) the reduction of the 700 hPa equatorial easterly jet (over South America and the eastern equatorial Pacific); (3) the anomalous Hadley cell circulation that sets in during El Niøo over tropical South America; (4) changes in the flow of atmospheric moisture into the continent; (5) changes in atmospheric pressures resulting in the displacement of the convection centers within the ITCZ to the west and south; (6) feedbacks between precipitation and surface convergence in tropical South America [72,92]; and (8) land-atmosphere interactions due to the regional coupling between soil moisture, precipitation, and vegetation [22, 80, 83, 84, 93]. The physical mechanisms explaining the climatic anomalies related to ENSO in Colombia have been studied by [83-86,90,94-99], among others. In the Pacific region the maximum correlation between ONI and precipitation is «0.65, while in the Amazon and Orinoco regions, correlations are not statistically significant. These results are in agreement with those found by Salas et al. [100].

The largest correlation between ONI and precipitation at national scale is simultaneous (lag 0) (see S1 Fig). Both temperatures exhibit a positive correlations with ONI in most of the country, with the exception of the Amazon region for *T_max_*, and higher correlations in the Caribbean region. Correlations between ONI and both temperatures get maximum values at varying lags throughout the country. The Pacific and Caribbean regions show higher correlations for *T_max_* at lag 0, while the Andes and Orinoco regions exhibit largest correlations at 1 to 3-month lags.

### Dengue in Colombia

Colombia has ecological, climatic, and entomological conditions that make suitable the survival of *Ae. aegypti* in the humid and hot low-lands (90%) of the territory [47,101,102]. Dengue infection rates show an increasing trend in the last decades [103-106]. The maximum altitude at which the *Ae. aegypti* is found has increased from 1,800 to 2,200 m a.s.l. between 2010 and 2013 [104,106], which can be attributed to climate change and deforestation [67,68,107,108]. The largest epidemic outbreak occurred during 2010 (see Fig 2), with around 150,000 cases, 9000 severe dengue, and 220 deaths [49,104], followed by 2013 and 2016. The departments with more cases historically have been Antioquia (Andean and Caribbean regions) and Valle del Cauca (Andean and Pacific regions) [104,106,109].

**Fig 2.**
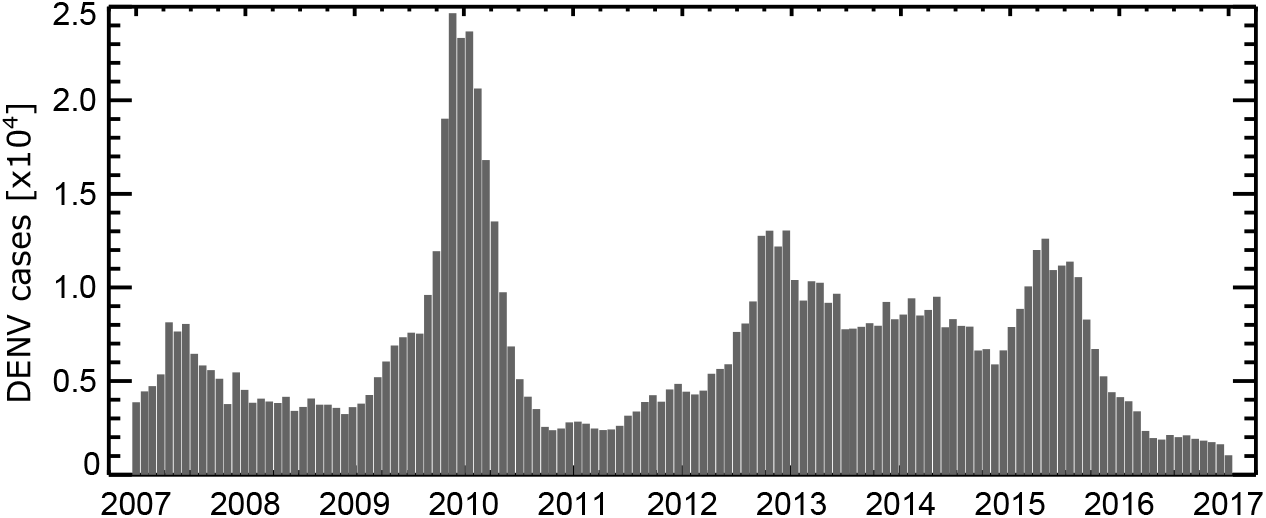
Time-series of dengue cases in Colombia. Monthly time-series of DENV cases.

Fig 3 shows the wavelet and Fourier spectra of ONI and DENV cases in Colombia. The largest portion of the variance of both variables are explained by 32-month periodicities, being stronger from 2009 to 2013, followed by a period of about 62 months, although it lies outside (in both cases) the significant cone of influence (non-hatched). Such periods evidence the strong association between dengue cases at ENSO frequencies. This association is studied throughout this paper. Besides, this figure also shows a large power around an 18-month period between 2009 and 2011 years. This time interval coincides with the largest number of dengue cases in Colombia that occurred during El Niøo 2009-10 and the transition to La Nina 2010-11.

**Fig 3.**
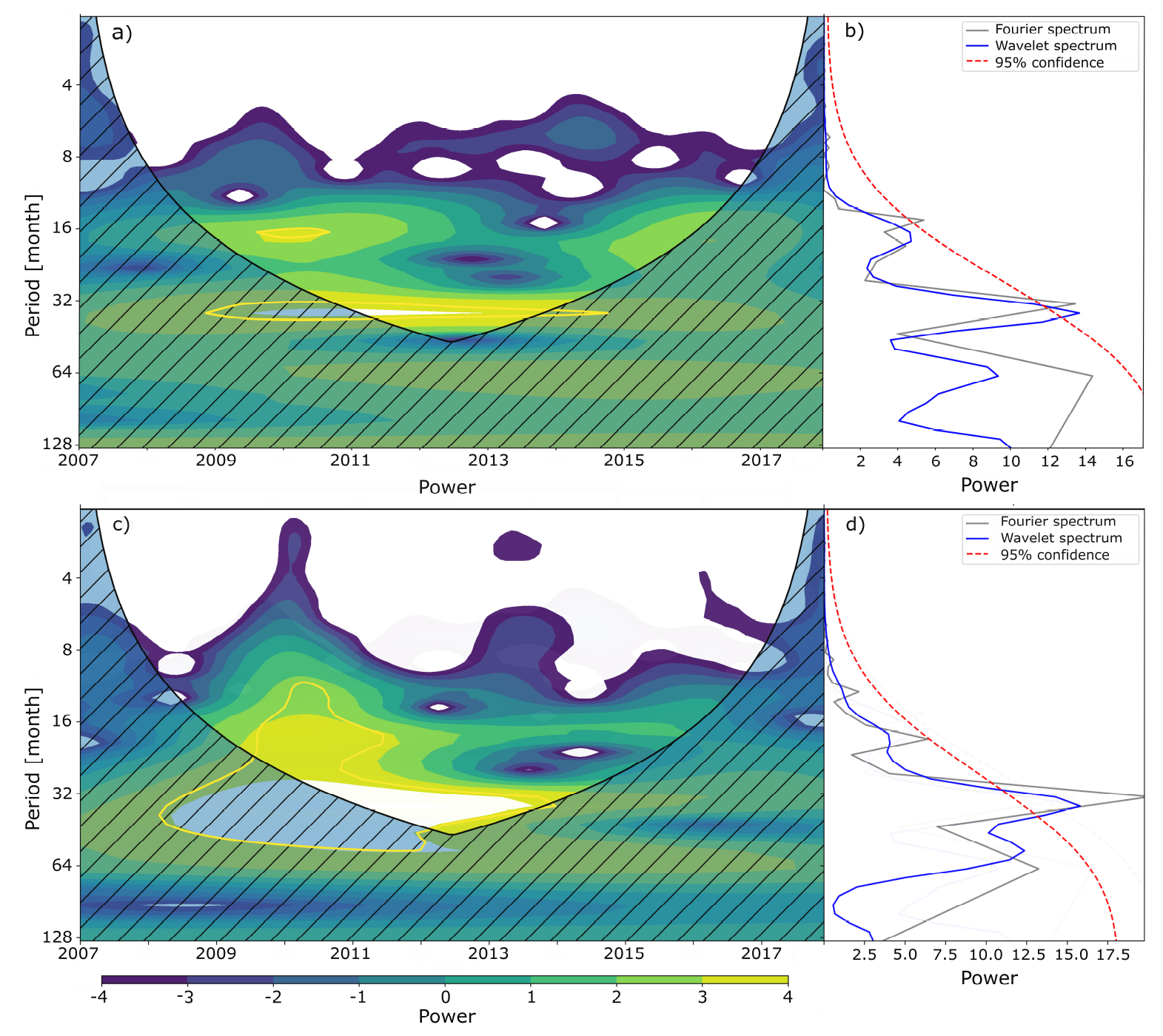
Wavelet analysis of ONI and DENV cases in Colombia. Wavelet power spectrum, global power spectrum and Fourier spectrum of ONI (a,b) and DENV cases (c,d).

### Linkages Between Local Climate, ENSO and Dengue

For our purposes, monthly dengue cases data and climate variables data are aggregated at national, regional, departmental, and municipal scales. At national scale, we use three methods to quantify the association between dengue cases, climate variables, and ENSO: lagged cross-correlations, cross-wavelet transforms, and the four variants of the nonlinear PCMCI causality (ParrCorr, CMI, GPDC, and PCMCIplus). Also, at national scale we estimate causality measures to quantify the nonlinear dependencies among dengue, ESPI, minimum and maximum temperatures, and precipitation. At smaller political scales, we only estimate cross-correlation due to the few data available. At national and regional scales, we also analyze the association between dengue cases and ONI at seasonal timescales.

#### National scale

Fig 4a shows the time-series of standardized monthly cases of dengue (left y-axis) and ONI (right y-axis) between 2007 and 2017. The red (blue) dashed line indicates the threshold of 0.5 (−0.5) °C, which are used to denote both phases of ENSO, as per the National Oceanic and Atmospheric Administration of USA (NOAA). The red (blue) polygons denote the occurrence of El Niño (La Niña), taking also into account the persistence of the ONI values above or below the threshold for more than 5 months. Fig 4a shows that an increase (decrease) of the ONI index (representing the warming of sea surface temperatures in the tropical Pacific) is followed by an increase (decrease) in the number of dengue cases in Colombia, especially after the 2009-10 El Niøo event. Of note is that the magnitude of the dengue peaks are not necessarily proportional to the magnitude of ENSO events given the multifactorial nature of dengue incidence that include socioeconomic conditions and the responsiveness of public health interventions to prevent and control the disease. The time-lagged cross-correlations shown in Fig 4b quantify the linear association between dengue cases and ONI, showing high positive correlations for lags between 2 and 7 months, and also suggest that El Niño (La Niña) events are associated with an increase (decrease) of dengue cases in Colombia with a 2 to 6-months lag.

**Fig 4.**
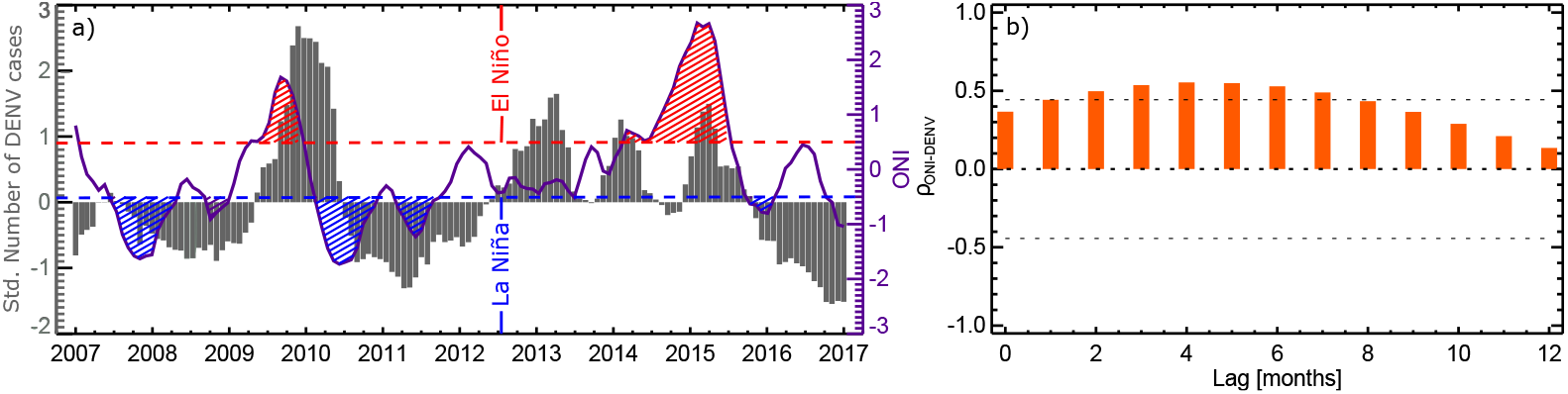
Time-series and cross-correlation of ONI and dengue cases in Colombia. Time-series (a) and cross-correlation (b) of ONI and dengue cases. Dashed lines in (a) indicate the threshold that determines the occurrence of El Niño (0.5) and La Niña (−0.5) according to NOAA, and in (b) the 95% confidence interval.

In spite of the previously shown high cross-correlations between ONI and dengue in Colombia, they are lower than those found by Acosta and co-authors [47] for the period spanning from 2005 to 2013. With the aim to understand this difference, we calculate the dynamic correlations between the ONI and dengue cases for lags from 0 to 5 months. These cross-correlations are estimated by aggregating the data from both series, one month at a time, after December 2009, that is:

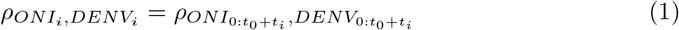

where *t_0_* is the December of 2009 and *t_i_* the time at *i*.

Fig 5 shows the dynamic cross-correlations, each line representing a time lag between 0 (darker blue) and 5 (lighter blue) months, indicating that the latter years of the study period exhibit lower correlations (from around 2015), and 2010 exhibits the highest ones.

**Fig 5.**
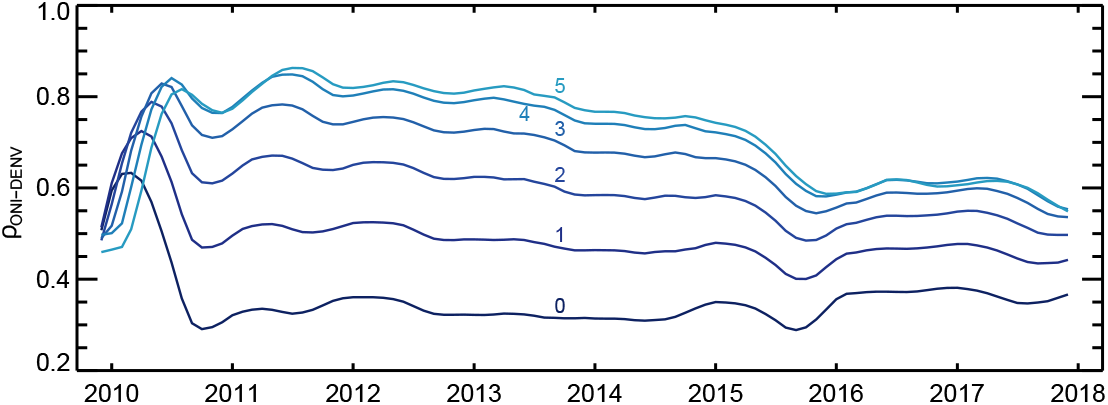
Dynamic cross-correlations between ONI and the number of dengue cases in Colombia. The different lines denote the lags between 0 and 5-months.

Note that 2010 is the year with more cases in Colombia during the study period. Since 2011, correlations at lag 0 show small changes, while at 3 to 5-month lags (higher correlations), correlations decreased markedly in recent years. This explains the differences found between the results of Acosta et al. [47] and Fig 4b.

**Fig 6.**
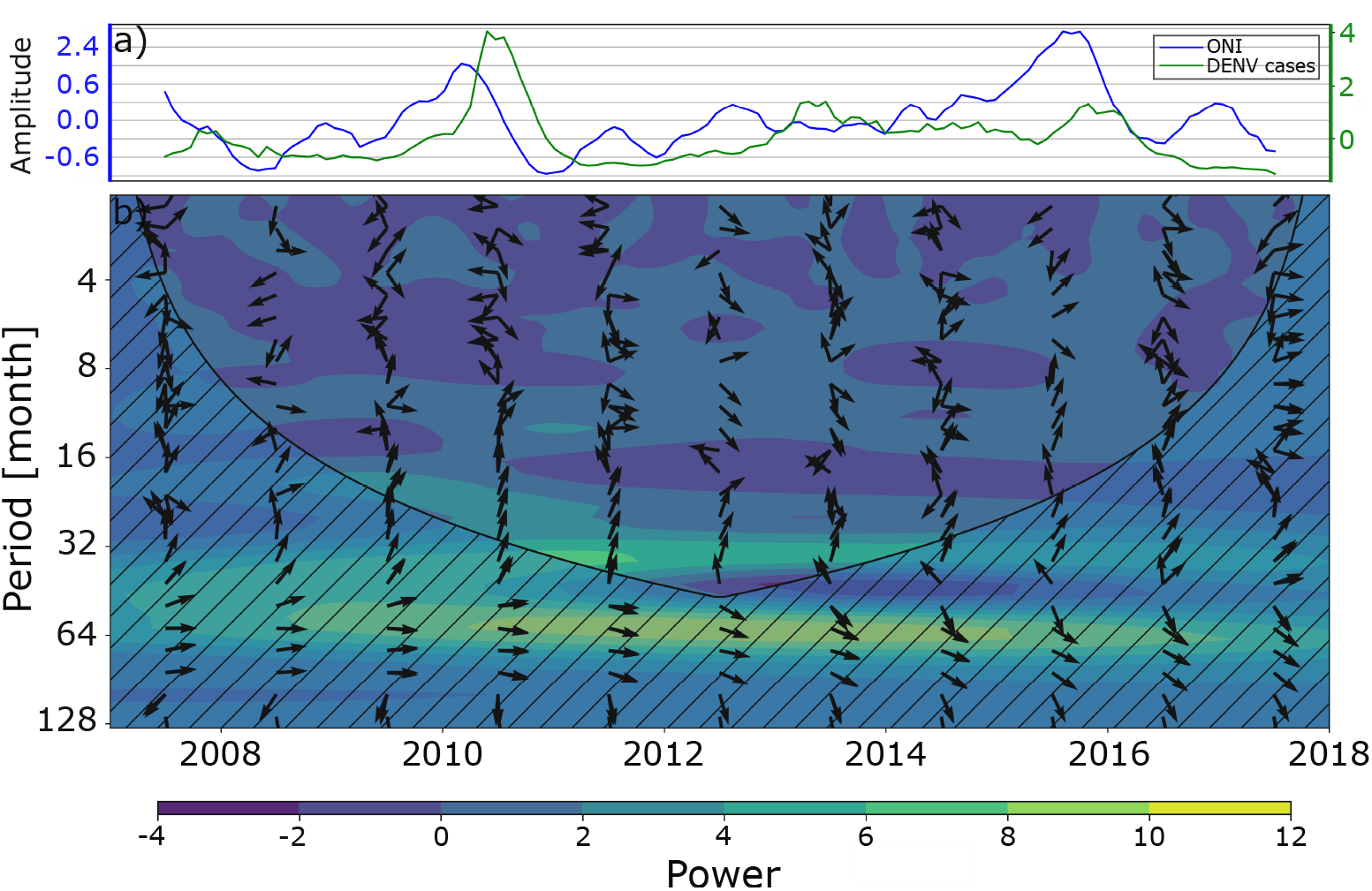
Cross-wavelet analysis between ONI and dengue cases in Colombia. Time-series of ONI and standardize cases of dengue used for the wavelet analysis (a) and cross wavelet transform (b). The relative phase relationship is shown as arrows with in-phase pointing right and anti-phase pointing left.

Fig 6 shows the cross-wavelet transform between ONI and dengue cases in Colombia, using a Morlet mother wavelet. The arrows indicate the relative phase relationship between the two variables, with in-phase pointing right and out-phase pointing left. The greatest power is observed at approximately 64 months period (≈ 5 years) for the entire record length. Both phenomena show an almost completely in-phase behavior until around 2011, and it goes losing this as time goes on, although this period is out of the statistically significant (non-hatched) cone of influence. Also, there is a high power near 32 months (≈ 2.5 years), although the variables are not fully in-phase. Both periods of 32 and 64 months are associated with the occurrence of the extreme phases of ENSO.

Fig 7 shows lagged seasonal (DJF, MAM, JJA, and SON) cross-correlations between ONI and raw series of dengue cases in Colombia. Here we use raw rather than standardized data to investigate seasonal differences of ENSO and climatic variables on dengue incidence. Purple bars correspond to correlations between ONI and dengue cases during the same season. To describe the results, we use brackets with two positions, where the first position indicates the quarter of ONI and the second the quarter of DENV cases. Higher cross-correlation appear for (DJF,DJF) and (SON,DJF), and lower for (MAM,DJF). SON is the season of ONI that results in higher correlations, and DJF is the season of dengue cases that seems to be more affected by ENSO (see [79,84])(except for ONI in MAM). Remarkably, the only negative cross-correlation occurs in (JJA,JJA), but it does not exceed the confidence interval of statistical significance.

**Fig 7.**
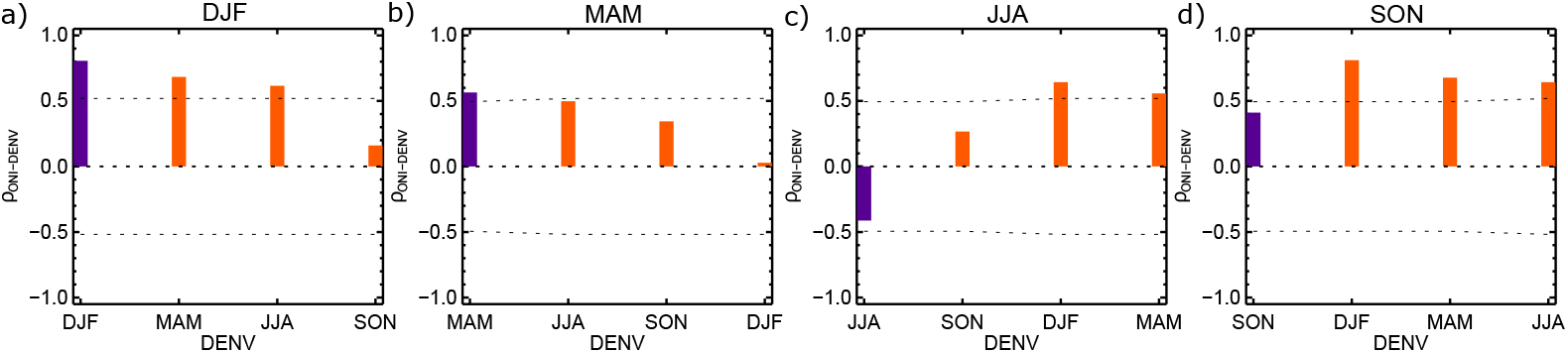
Seasonal lagged cross-correlation between ONI and dengue cases. During DJF (a), MAM (b), JJA (c), and SON (d). Dashed lines indicate the 95% confidence interval.

As mentioned before, ENSO does not affect directly the dynamics of dengue dynamics, but modify weather and climate variables that are directly related to dengue transmission. To identify which variables affected by ENSO influence dengue in Colombia, we performed cross-correlation analysis between the number of DENV cases and precipitation, maximum and minimum temperature, wind velocity, and relative humidity. Table 1 shows the maximum cross-correlations between climate variables and dengue cases and the peak lag associated with them. Unlike other sites (e.g., [8,27,110,111]), precipitation is negatively correlated with the number of dengue cases in Colombia. Maximum temperature is highly and positively correlated with dengue cases as expected, and wind speed is positively related as it can help the spread of mosquitoes. Finally, relative humidity is negatively correlated to DENV cases (even if correlations do not exceed the CI), suggesting that the water stress effect is important in Colombia. Confronting these results with those in Fig 4, it is possible to suspect that the key variable in the relationship between ENSO and dengue cases is temperature, as noted in other sites (e.g., [22,27,67,112-114]).

**Table 1.** Maximum correlations and lags between climatic variables and dengue cases on the national scale.

| Variable | Max. $\rho$ | lag [month] |
| --- | --- | --- |
| $P$ | -0.532 | 6 |
| $T_{max}$ | 0.551 | 5 |
| $T_{min}$ | 0.300* | 3 |
| $WV$ | 0.468 | 4 |
| $HR$ | -0.415* | 6 |

\*This value does not exceed the confidence interval.

Now we focus on results about non-linear causality metrics based on PCMCI. We use *α*=0.01 and *t_max_* = 9 months. Figs 8 and 9 show the results of the causal inference methods taking into consideration the maximum and minimum temperature, respectively. ParCorr indicates positive causality between ESPI and DENV cases (for lags between 1 and 9 months), while the PCMCIplus method indicates negative causality. Positive causalities point out that increases in SST over the Pacific Ocean are non-linearly associated to increases in the number of dengue cases. The PCMIplus method shows an indirect relationship between ESPI and DENV cases when *T_max_* is taken into account and no relations when *T_min_* does. Both methods show a negative relationship between ESPI and P, for lags between 1 and 3 months.

**Fig 8.**
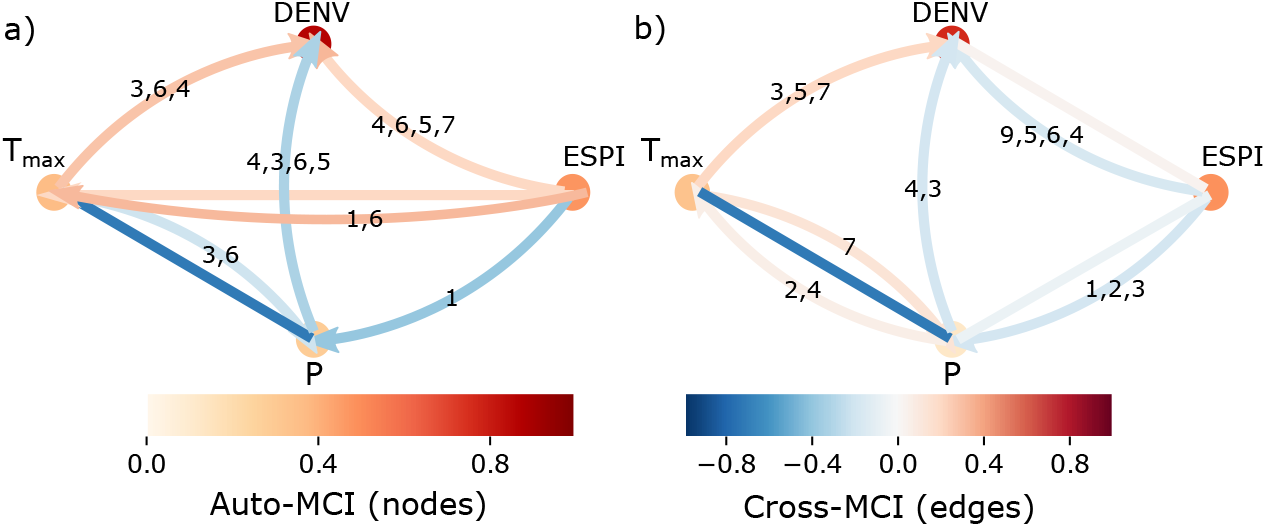
Non-linear causalities between DENV cases, ESPI, *P*, and *T_max_* in Colombia. Non-linear causalities given by ParCorr (a), and PCMCIplus (b) for data.

**Fig 9.**
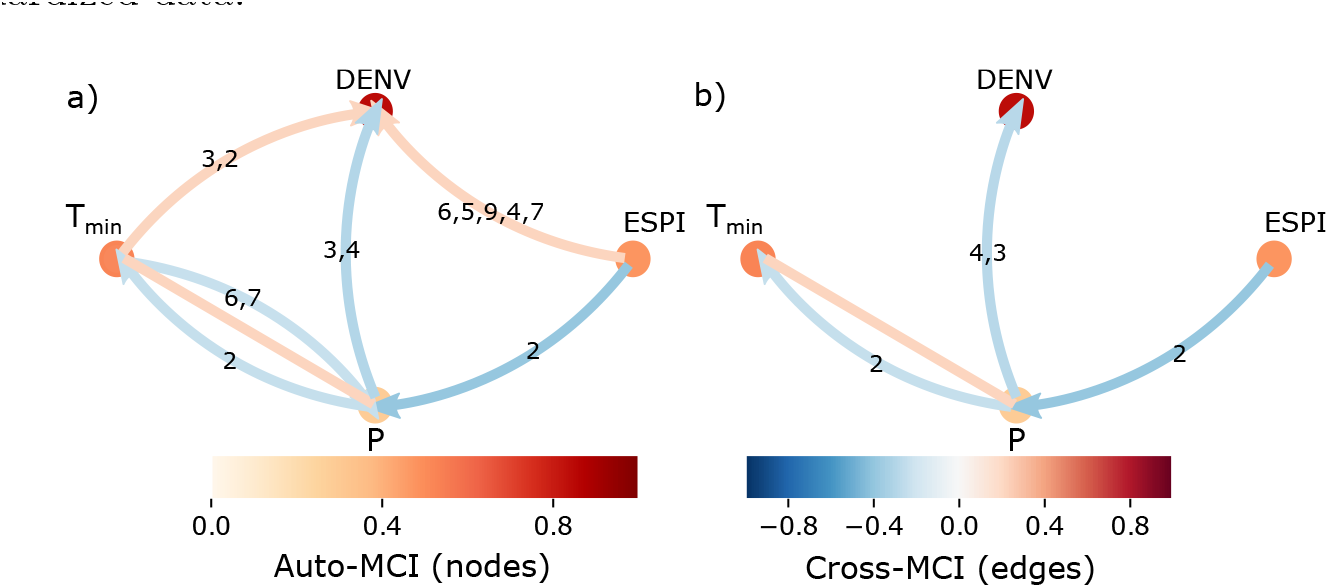
Non-linear causalities between DENV cases, ESPI, *P*, and *T_max_*, in Colombia. Non-linear causalities given by ParCorr (a), and PCMCIplus (b) for standardized data.

Results from the ParCorr test indicate positive relations between ESPI and *T_max_* for lags of 1 and 6 months and 1 month, respectively. PCMCIplus shows no association but indicates that ENSO modifies precipitation and this, in turn, changes temperature (for lags between 4 and 7 months). The ParrCorr method also indicates that the effects of ENSO has on *T_min_*, if they occur, are brought about by precipitation. The non-linear causalities between P and both temperatures *(T_max_* and *T_min_)* are negatives for the ParrCorr method (for *T_max_* the causality is indirect, and for *T_min_* it occurs for a lag of 2 months). PCMCIplus shows a positive causality of P with *T_max_* and a negative with *T_min_* (for lags of 2 and 4 months for *T_max_* and of 2 months for *T_min_)*. The PCMCIplus method shows an indirect causality with *T*. Negative and positive relationships are physically possible, negative ones are the result of higher evapotranspiration *(ET*) rates given the increase in solar radiation and more water in the soil. High *ET* rates mean more energy converted into latent heat, decreasing the flux of sensible heat. This can lead a loop since when *ET* increases, local rainfall increases, and the same happens with soil moisture (s). In turn, high values of s (in combination with the states of other variables such as available radiation, relative humidity, CO_2_, etc.) result in high values of ET. On the other hand, positive causalities can occur because, after a decrease of *T*, *ET* can also decrease due to a decrease in water vapor deficit, thus decreasing local precipitation, and then *ET* [115].

ParCorr shows negative non-linear causalities between T and P (for lags of 3 and 6 months for *T_max_* and 6 and 7 months for *T_min_*), and PCMCIplus indicates a positive relation of *T_max_* and P for a lag of 7 months and a negative indirect relationship. Non-linear causalities between precipitation and DENV cases are negative for all methods for lags between 3 and 6 months. Both negative and positive causalities are found worldwide as discussed in Section Discussion.

All methods show positive causalities between *T_max_* and DENV cases for lags between 1 and 7 months. This association has been identified in many sites, however, it is not direct and depends on the temperatures range. Furthermore, ParCorr indicates positive causalities between *T_min_* and DENV cases for lags between 2 and 3 months, but the PCMCIplus method shows no causality, suggesting that minimum temperature only affects through precipitation, with which it has a positive indirect association.

Note that, as expected, all results obtained with the non-linear methods do not show any causality between local climatic variables on ESPI, DENV cases on ESPI, and DENV cases on the climatic variables.

#### Regional scale

Fig 10 shows regional maps of maximum correlations, *ρ_max_*, and corresponding lags between ONI and dengue cases. Only the Pacific and Andes regions exhibit very high statistically significant cross-correlations. The highest cross-correlations appear in the Pacific region, showing values around 0.7 and the lowest ones in the Caribbean region with values close to 0.2. Higher correlations in the Orinoco region are negative, but as mentioned before, they do not exceed the CI. The highest correlation in the Pacific region occurs at 4-month lag, while at 6-months lag in the Andes region, which indicates a delay in the effects of ENSO in the latter region respect to the former one. Here, it is possible to observe the strong influence of the Pacific and Andes regions on the overall effect of ENSO in dengue cases at national scale since correlations continue to be positive and very high (*ρ* ≈ 0.55), and the associated lag is the same as that of the Pacific region, i.e., 4 months. The maximum correlation value at the national level is lower than that of the Pacific region due to the lower cross-correlations in the Caribbean, Orinoco and Amazon regions. However, these latter regions do not get to remove the relevance of ENSO’s effect on dengue dynamics on the aggregated national scale.

**Fig 10.**
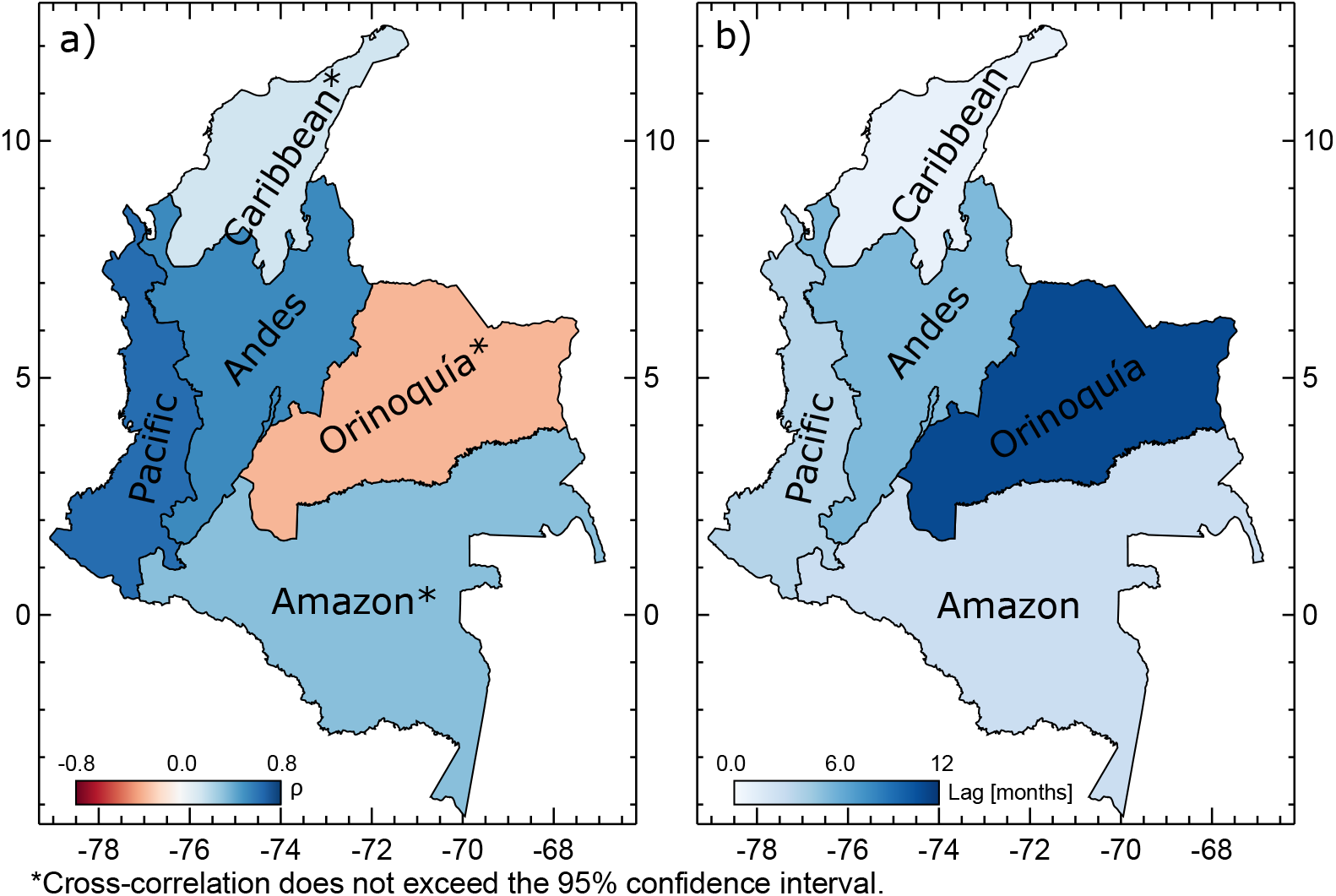
Cross-correlations between ONI and DENV cases at the regional scale. Maximum cross-correlation (a) and associated lag (b) between ONI and DENV cases.

Fig 11 shows non-linear causality results for all regions. As we noted in the previous analyzes, the nationally-aggregated behavior is mainly affected the situation in the Pacific and Andes regions, especially the first one. Precipitation is affected by ENSO in the Andes and Pacific regions according to the ParCorr and PCMCIplus methods, and in the Orinoco and Caribbean regions according to the ParCorr method. The effects in the Andes, Caribbean, and Pacific are at 1-month lag, while in Orinoco at 7 and 5-months lags. The higher cross-MCI occurs in the Andes region.

**Fig 11.**
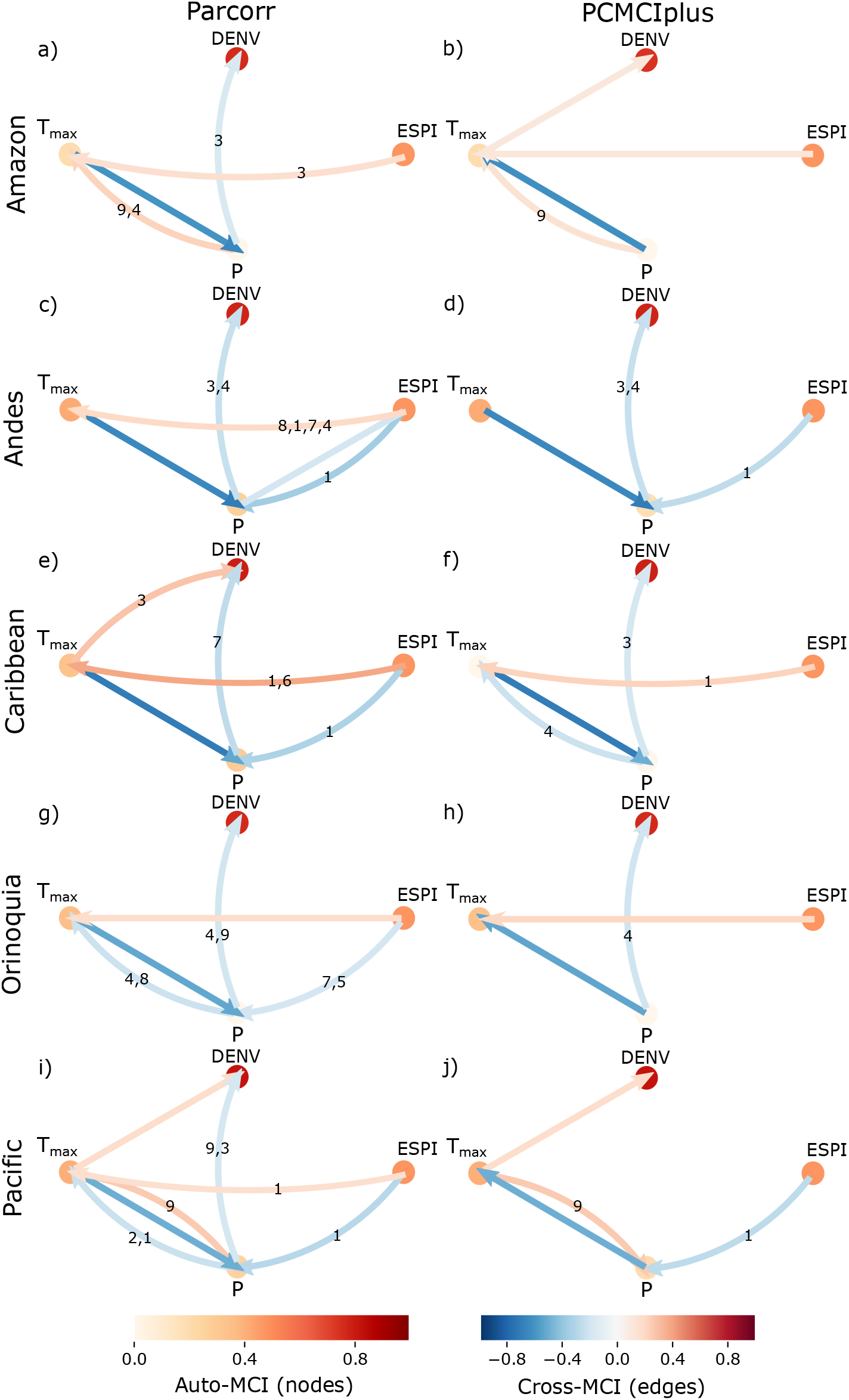
Non-linear causalities between DENV cases, ESPI, P the regional scale at regional scale. Non-linear causalities given by ParCorr (left panel), and PCMCIplus (rigth panel) for standardized data.

ENSO modifies maximum temperature in the Amazon, Caribbean and Orinoco regions with both methods (in the Orinoco region the relationship is indirect), and in the Andes and Pacific it only appears with the ParCorr method. These associations are for lags between 1 and 8 months, with highest values in the Caribbean and Andes regions. Precipitation and maximum temperature are related in all regions but only in the Caribbean and Pacific regions show two-directional relationships with the PCMCIplus method.

Precipitation shows a direct causality on the number of DENV cases in all regions with the ParCorr method, and in the Andes, Caribbean and Orinoco regions with the PCMCIplus method. In these two latter regions, precipitation can affect DENV cases through changes in temperature. On the other hand, *T_max_* only directly affects dengue cases in the Caribbean region for a lag of 3 months with the ParCorr method. Dengue cases in the Pacific and Amazon regions are only indirectly affected by *T_max_*. If in the other regions, there is any effect of *T_max_*, it must be through the precipitation, as in Amazon and Orinoquia with the ParCorr method.

Fig 12 shows lagged seasonal cross-correlations maps, indicating that the effects of ENSO vary in space and time. Higher cross-correlations between (ONI, Dengue) occur during (DJF, DJF), (DJF, MAM), (MAM, MAM), (JJA, DJF), (JJA, MAM), (SON, DJF), (SON, MAM), and (SON,JJA), especially in the Pacific and Andes regions.

**Fig 12.**
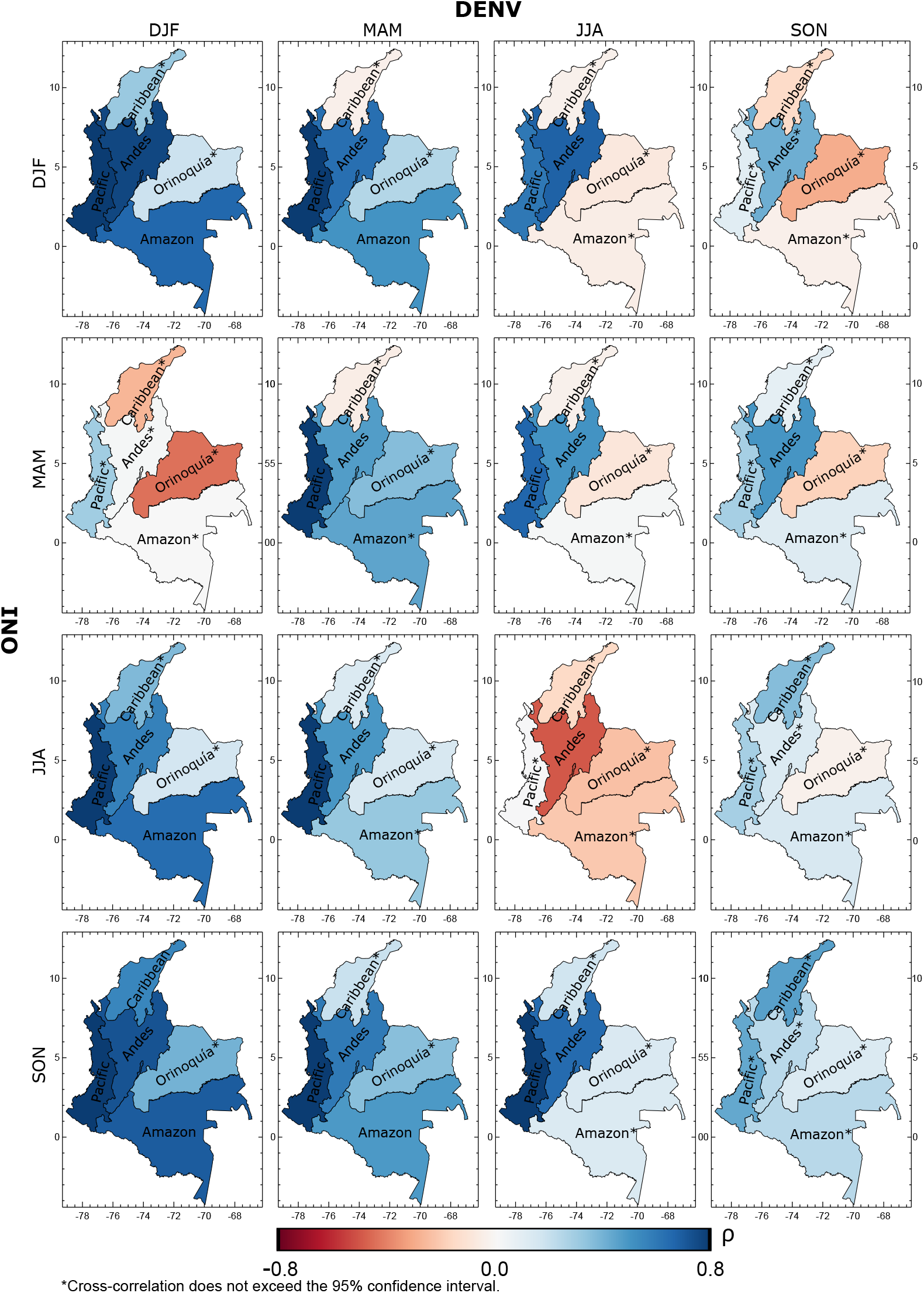
Regional seasonal cross-correlation maps between ONI and DENV cases.

The association of ENSO with dengue cases during some seasons is remarkably strong in the Amazon region, such as in (DJF, DJF), (DJF, MAM), (JJA, DJF), (SON, DJF), and (SON, MAM). Correlations in the Caribbean and Orinoco regions continue to be very low, except in Orinoco during (MAM, DJF), on the order of -0.44, but in no case they exceed the CI. Results at the national scale only show negative correlations for (JJA, JJA) (see Fig 7). This season continues to exhibit the most notable negative correlations at the regional scale, although only the correlation of the Andes exceeds the CI. Orinoco, Amazon, and Caribbean also show negative correlations in other quarterly combinations, e.g. (DEF, JJA) and (DJF, SON), but none of them exceed the CI. The only quarterly combination in which the Andes shows negative correlations (−0.5) is (JJA, JJA), indicating again the strong influence of this region at the national scale. Also, the lowest correlations at the national scale appear in (MAM, DJF) as in the Andes region.

Table 2 shows results of maximum correlations, *ρ_max_*s, between climatic variables and dengue cases and the corresponding lag at regional scales. The linear association between dengue cases and precipitation is negative in the Andes and Pacific regions, like the national scale. On the other hand, maximum temperature shows high and positive correlations with dengue in the Andes and Pacific regions, especially in the latter, and minimum temperature in the Pacific region. Although the correlation values do not exceed the confidence interval in the Orinoco region, the negative correlations between DENV cases and *T_min_* can explain the negative association between ONI and DENV in this region. Similarly, the negative associations between P and DENV and the positive associations between temperature and DENV would explain the high positive correlations between ONI and DENV in Andes and Pacific regions. The mechanistic processes involved in these associations are discussed in Section Discussion.

**Table 2.** Maximum cross-correlations and lags between climatic variables and dengue cases for the different regions.

| Region | $P$ | | $T_{max}$ | | $T_{min}$ | |
| --- | --- | --- | --- | --- | --- | --- |
| | Max. $\rho$ | Lag | Max. $\rho$ | Lag | Max. $\rho$ | Lag |
| Amazon | 0.210* | 0 | 0.207* | 5 | 0.330* | 1 |
| Andes | -0.511 | 6 | 0.422 | 6 | -0.193* | 12 |
| Caribbean | -0.295* | 4 | 0.289* | 4 | 0.306* | 0 |
| Orinoco | -0.160* | 5 | -0.196* | 12 | -0.404* | 12 |
| Pacific | -0.505 | 5 | 0.624 | 1 | 0.473 | 1 |

\*This value does not exceed the confidence interval.

#### Departmental scale

Similar analysis at the department scale allow us to conclude that the association between ONI and DENV cases in each region is determined by a few departments, as shown in Fig 13. Antioquia (Andes and Caribbean regions) exhibits higher correlations, followed by Valle del Cauca (Pacific and Andes regions). As mentioned above, these are the departments with the highest number of cases historically. The departments with higher correlations are located, in general, over the Andes regions (low-lands and piedmonts). Fig 13b shows the lags associated with highest correlations in each department, pointing higher lags for departments with negative but not statistically significant correlations. Departments located near the Pacific Ocean (western zone) exhibit smaller time lags, as on the regional scale.

**Fig 13.**
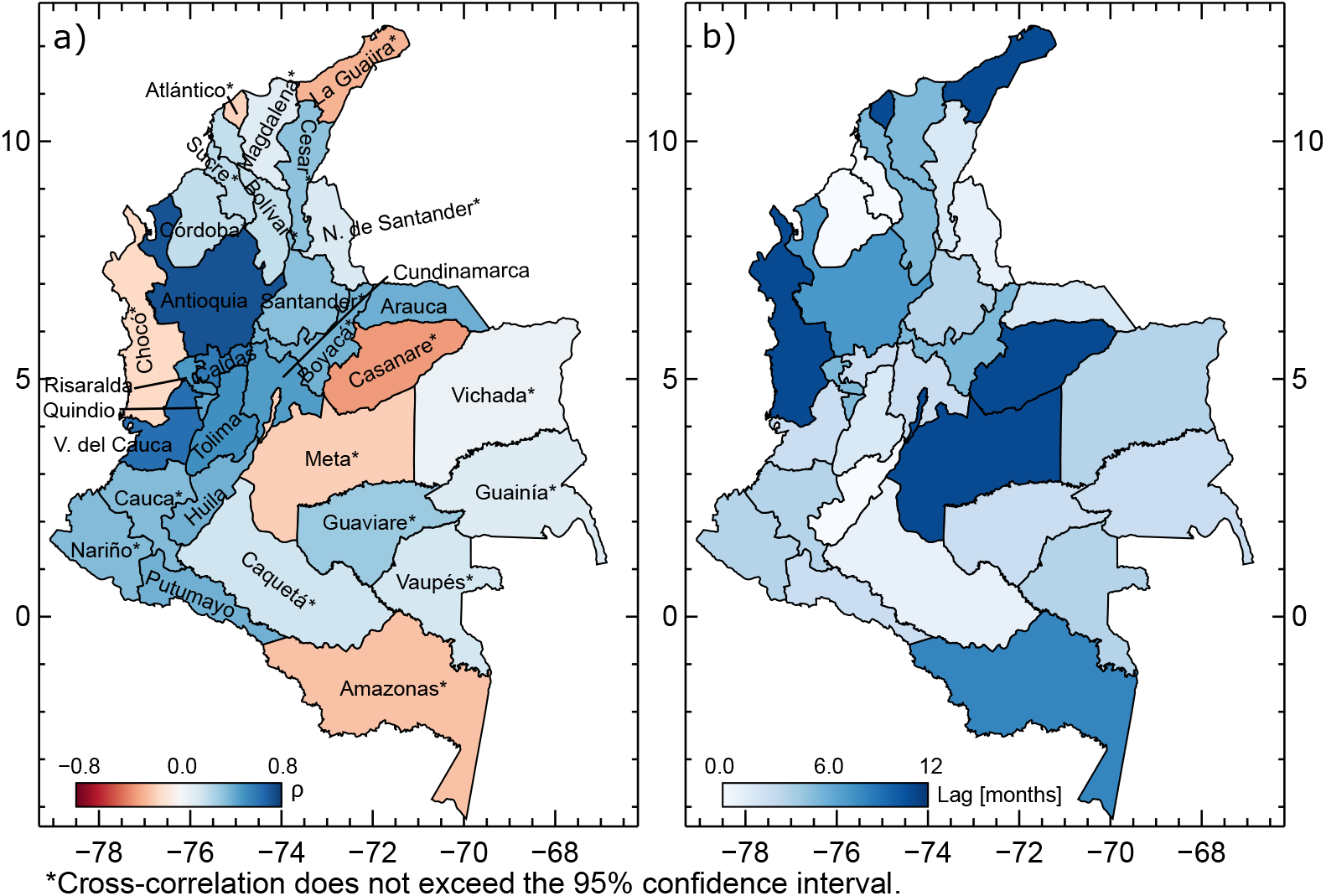
Cross-correlation between ONI and dengue cases at the departmental scale. (a) Maximum cross-correlation and (b) associated lag between ONI and DENV cases.

Fig 14 shows the maximum correlations estimated between climatic variables and DENV cases at departmental scale. The highest correlations with precipitation and maximum temperature appear in Antioquia and Valle del Cauca (as *ρ_max_* of ONI and DENV cases), followed by Caldas, Tolima, and Quindio. P and dengue cases exhibit negative correlations in almost all the departments of Colombia, while in the rest do not exceed the CI. *T_max_* and dengue cases are positively correlated in all the departments showing a statistically significant result, except in Putumayo (Amazon region). *T_min_* shows the highest correlations at Tolima (Andes region), and negative correlations at Cundinamarca (Andes region) and Putumayo. These latter departments show positive correlations between ONI and DENV cases, which suggests that in Cundinamarca, ENSO-driven variations in temperature do not influence the dynamics of dengue but changes in the precipitation. In Putumayo, dengue cases are not significantly correlated with precipitation, and maximum and minimum temperatures show negative correlations, indicating that a decrease in temperature could lead to an increase in dengue cases. However, ENSO has a slight effect in this zone (see Fig 1).

**Fig 14.**
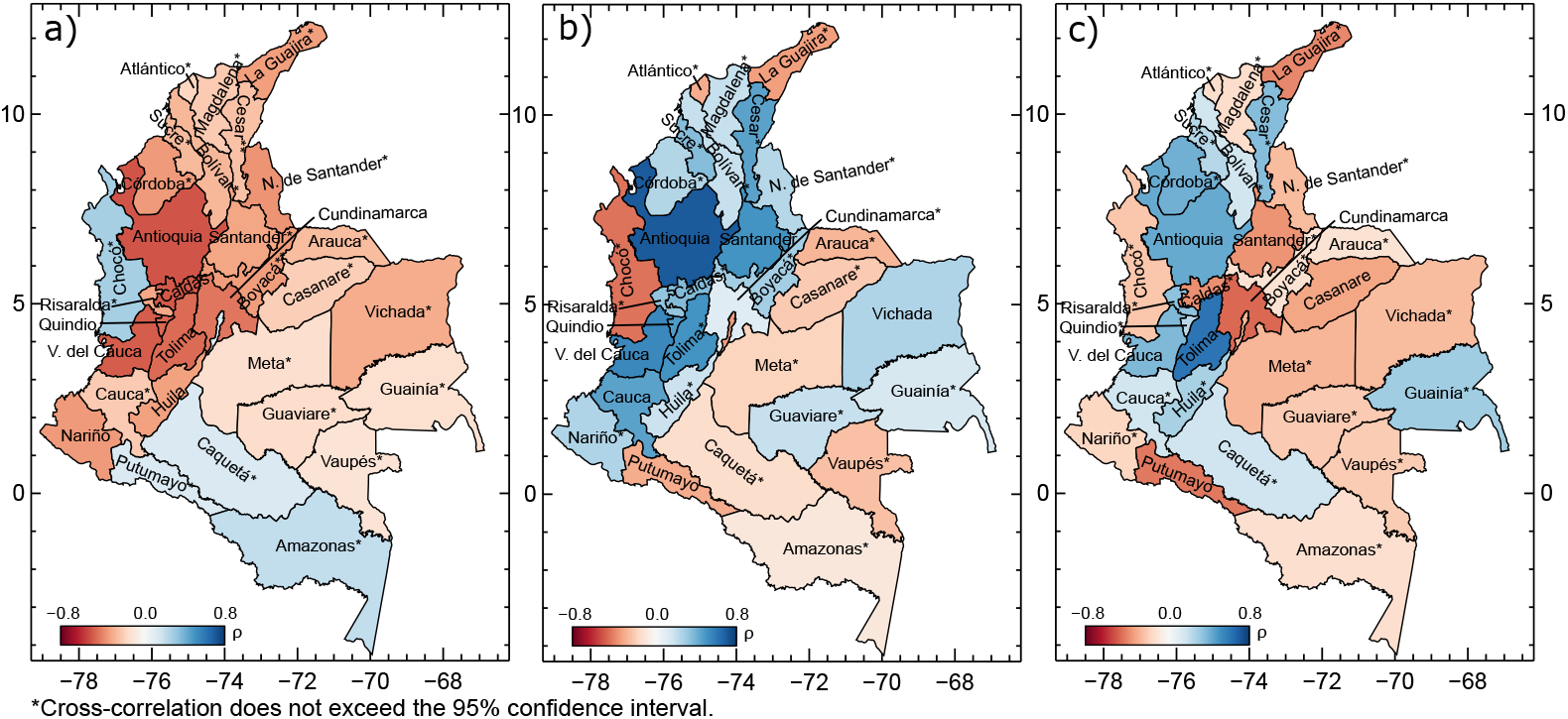
Cross-correlation between dengue cases and climate variables at the departmental scale. Maximum cross-correlations between DENV cases and precipitation (a), maximum temperature (b), and minimum temperature (c).

#### Municipal scale

One step down in the spatial scale, in most cases it is was not possible to estimate cross-correlograms due to lack of long enough data. Here, we select the two largest cities with the highest number of cases both in the analyzed period and historically, i.e., Medellín (Antioquia department, Andes and Caribbean regions) and Cali (Valle del

Cauca department, Pacific and Andes regions). Fig 15 shows the time series of the standardized number of dengue cases and ONI and their cross-correlograms. Both cities show a significant increase in the number of cases after the 2009-10 and 2015-16 El Niøo events. Besides, Cali exhibited an increase in dengue after the warming of the Pacific Ocean during 2011-2012, although it was not classified by NOAA as an El Niño event. Cross-correlograms show very high values, particularly in Medellín (*p_max_* ≈0.8). When comparing the lags associated with higher correlations in both cities, it is possible to observe similar delays as in the the regional and departmental scales. The effects of ENSO in Cali start much earlier (indeed they can be simultaneous) while in Medellín they occur after 3-4 months, with the highest correlations at 7-months lag.

**Fig 15.**
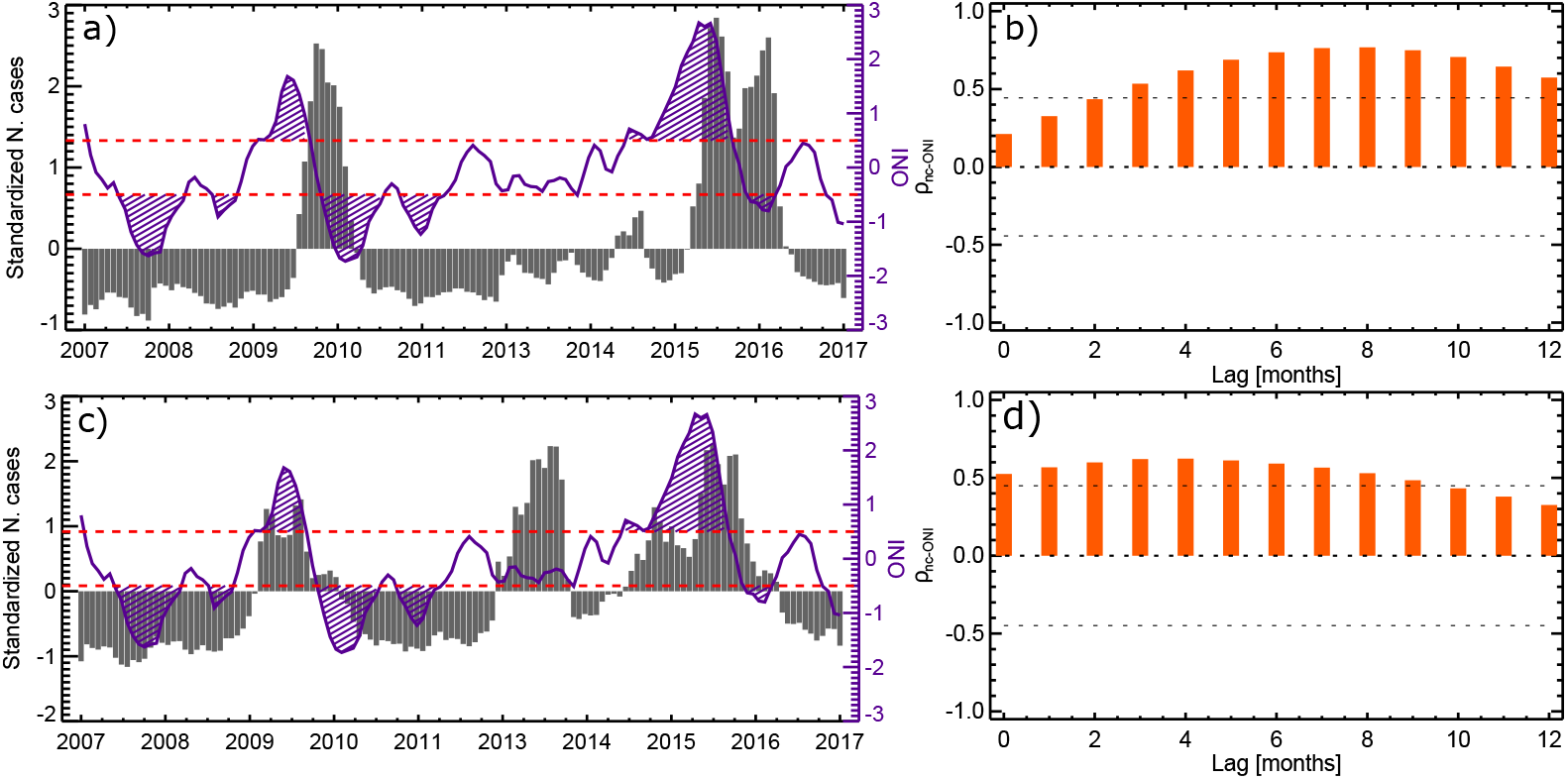
Time-series and cross-correlogram between ONI and dengue cases at the local scale. Time-series and cross-correlogram of ONI and dengue cases of Medellín (a,b), and Cali (c,d). Dashed lines in (a) and (c) indicate the occurrence of El Niøo and La Nina, and in (b) and (d) indicate the 95 % confidence interval.

Table 3 shows the cross-correlation at lag 0 between the three climatic variables and the number of DENV cases. In Medellín, no correlation exceeds the confidence interval, but the signs remain the same as for department of Antioquia. In Cali, the correlation with precipitation does not exceed the CI either, but the association with minimum and maximum temperatures seems to play an important role, especially with the former one.

**Table 3.** Cross-correlation between climatic variables and dengue cases in Medellín and Cali at lag 0.

| Variable | Medellín | Cali |
| --- | --- | --- |
| $P$ | -0.1* | -0.26* |
| $T_{max}$ | 0.26* | 0.54 |
| $T_{min}$ | 0.36* | 0.57 |

\*This value does not exceed the confidence interval.

## Discussion

Our results confirm the strong relationship between ENSO and the incidence of arboviruses like dengue in many parts of the world [11,22,43,47,70,72]. Analyzes carried out in this study provide further evidences about the important association between ENSO, local climate and dengue dynamics in Colombia at a wide range ofspatial and political scales, unlike the non-statistically significant relationship found by [116] and in agreement with other previous studies [22,37,47]. El Niøo (La Nina) events are associated with the increase (decrease) in dengue cases in the country. Detailed analysis at finer spatial and temporal resolutions allowed us to identify regional, departmental and municipal differences, as well as within the year (see Figs 4b, 8, 9, 10a, 11, and 13a). Furthermore, we have identified that the association is not stationary, pointing to a reduction in dengue cases in Colombia since 2015 (see Fig 5), as in the case of the study by Cazelles et al. [11] in Thailand.

The highest (and positive) cross-correlations between ONI and DENV cases appear in the Pacific and Andes regions, while negative in the Orinoco region although below the CI (see Fig 10a). Note that as the Ae. Aegypti is tipically an urban mosquito [117] and the majority of Colombia’s population is concentrated in the Pacific, Andes, and Caribbean regions, most of cases belong to these regions. High cross-correlations in the Andes region are explained by the dynamics at the departments of Antioquia, Caldas, Risaralda, Tolima, and Cundinamarca. In the Andes region, all departments show varying albeit positive cross-correlations. The Pacific region exhibits the highest cross-correlations, mainly explained by the situation in the department of Valle del Cauca. Similar results were observed in the Orinoco, Caribbean and Amazon regions (see Fig 13a).

Regarding results between ENSO and dengue cases at regional scales, the Pacific region has shorter lags associated with higher correlations than the Andes region (see Fig 10b), confirming the traveling wave found by Acosta [47], although spanning a broader geographical setting. This traveling wave is not so clear in the non-linear causality methods since several relationships are indirect and have no lag associated. The ParCorr method indicates that when the maximum temperature is taken into account, the effects of ENSO have a longer delay in the Andes than in the Pacific region (see Fig 11). The presence of such delayed effect of ENSO on the occurrence of dengue at departmental level can be used as a pro-time to take preventive measures before potential outbreaks. In Orinoquia the correlation between ENSO and DENV cases is negative correlations (El Niøo (La Nina) is related to less (more) cases of dengue) and associated with the highest lag (12 months), but it is not statistically significant (*ρ* =-0.27).

Additionally, our results at seasonal timescales evidence that the highest cross-correlations between ONI and DENV cases appear during (DJF, DJF) and (DJF, SON) and the lowest at (MAM, DJF). The highest (lowest) simultaneous correlation occurs when ONI shows the steepest positive (negative) gradient, i.e., DJF (JJA), going from values of around -0.22 (≈0.03) to ≈0.0 (≈-00.5) (see S2 Fig). The higher correlation occurs during the quarters with the lowest values of both ONI and DENV cases, and the lower correlation during quarters with high values of ONI and low values of DENV cases. Besides, in the latter case, there is a 9-month lag, which may indicate that the effect of SST anomalies in the tropical Pacific in MAM has already been lost in DJF. The national annual cycles of cases of dengue and ONI (see S2 Fig) are in phase throughout most of the year. There are some differences in SON as DENV cases increase slightly in October while, the ONI decreases. Hyper-endemic years (2010, 2013, and 2016) have more pronounced annual cycles, in association with the annual cycle of the ONI.

Results at the national scale are consistent with those at regional scales (see Figs 7 and 12). Again, it is worth noticing the strong influence of the Andes and Pacific regions at the national scale, since the combination of seasons showing higher cross-correlations in these regions dominate the aggregated behavior. The above is also suggested by the results obtained with the non-linear causality metrics, especially the influence of the Pacific region at national scale (see Fig 11).

The association between El Niño occurrence and outbreaks of DENV cases can be explained in terms of an increase in temperatures (minimum and maximum) and a decrease in rainfall, which may favor the ecological, biological, and entomological components related to this disease [22]. Other variables such as wind velocity and relative humidity may play an important role in dengue dynamics, but the available data is not enough to perform analyzes below the national scale. Wind velocity shows a positive correlation with dengue cases, while relative humidity is negatively correlated, but it does no exceed the CI (see Table 1).

Precipitation is negatively correlated with dengue cases in most of the country, except in the Amazon region (although it does not exceed the CI), and in the departments of Chocó, Caquetá, Amazonas, and Putumayo (see Tables 1 and 2, and Fig 14). Linear correlations between *P* and DENV cases result at national and regional scales are in agreement with those found with the non-linear causality methods (see Figs 8 and 11), which may indicate that these correlations imply causation. These relationships would be expected to be positive because precipitation provides favorable habitats for the aquatic stages of the mosquito development, influencing the life cycle and distribution of the mosquito [67]. If so, the amount of mosquitoes and dengue cases should increase (decrease) during La Niña as in Hawaii [110]. In some places, precipitation is only important when breeding sites are not handmade [27] or in wetter areas, where rainfall is not a limiting-transmission factor [13,20,79]. However, in Colombia, the decrease in rainfall brought about by El Ninño is mostly associated with increases in the number of dengue cases, so decreased rainfall can lead to the formation of stagnant ponds [22,72]. A similar result was found by Poveda et al. [79] for malaria in Colombia and Juliano et al. [118] for dengue in Florida, USA.

Maximum temperature is positively correlated at the national level and in all regions but Orinoco (although below the CI). At the departmental scale, this is only true over the Andes mountains and in Vichada (Orinoco), Guaviare (Amazon), and Guainía (Amazon) (see Tables 1 and 2, and Fig 14). Results from the non-linear causality metrics demonstrate that the associations shown by the cross-correlations not always indicate causality. The PCMCIplus method does not indicate any direct causality between *T_max_* and DENV cases in the regions, but only at the national scale. The ParCorr method indicates a direct relation only in the Caribbean region, and indirect in the Amazon and Pacific (see Figs 8 and 11). The above suggests that in most sites there is no direct effect of maximum temperature, but it causes changes in other variables (such as precipitation, relative humidity, wind velocity, etc.), which in turn affect the dynamics of dengue.

Minimum temperature is also positively correlated with dengue cases nationally and in the Amazon, Caribbean, and Pacific regions. However, at the departmental scale, there are no clear spatial patterns. Temperature may be a key component in DENV dynamics due to its numerous interactions with other environmental and climatic factors [67,72,113,114]. High temperatures result in larger virus replication rates, less time of mosquito incubation and gonotrophic cycles, more frequent biting rates, and greater survival at all stages within the observed temperature values [18,22,67,112,114]. This means more time available for the mosquito reproduction and transmission of the virus [72], and a greater basic reproductive number [114],*R*_0_, defined as the expected number of cases directly generated by one case in a population where all individuals are susceptible to infection.

Focks et al. [114] concluded that under tropical conditions, temperature does not affect adult mosquito abundance, but the quantity of water-holding containers and the amount of available food for larval survival. However, extremely high temperatures can contribute to the reduction of oviposition sites by evaporation [27], lifespan reduction, and the inability of mosquitoes to feed and fly [18,21]. These facts can help to explain the low correlations found in the Caribbean region since temperatures are always high owing to low altitude, which are even higher during the El Niøo (see Figs 1b,c). On the other hand, when temperature is very low (below 18° C according to [113]), the oviposition abruptly diminishes. This implies that the effect of temperature on dengue dynamics is non-linear and depends on local conditions. Furthermore, not only the average or instantaneous temperature values can influence this phenomenon, but also their variability and fluctuations, which have not been considered in this study, as it occurs in other sites [40, 111].

Almeida et al. [13] and Dibo et al. [19] suggest that air humidity is one of the most significant climatic variables in determining Ae. Aegypti abundance. In general, positive correlations are expected between relative humidity and dengue cases, i.e., that low values of relative humidity result in fewer dengue cases, given that the mosquito survival and egg development decline [67], and mosquitoes use their available cellular resources for their maintenance and not the virus [18]. At the same time, low humidity can place mosquitoes under stress, impeding them from fighting off a viral infection [18]. Besides, dehydration may cause an increase in blood feeding (biting rates), as female mosquitoes seek to ensure their reproduction [119]. Atmospheric humidity also influences evapotranspiration rates [114] and, as mentioned before, it may or may not favor the creation of breeding sites. Although humidity data is limited and the estimated correlations with dengue cases are not statistically significant, the value surpasses -0.41 (see Table 1). This suggests that the increase in blood feeding due to dehydration plays an important role in Colombia. Strong winds can extend the mosquito’s flight distance, but it can also reduce the biting frequency [61]. In Colombia, the first mechanism seems to have a greater influence, since dengue cases show a statistically significant and positive correlation with wind velocity (see Table 1). Notably, temperature, relative humidity, wind speed, and precipitation are not independent variables, with multiple nonlinear feedbacks among them.

Dengue transmission is a multi-factorial phenomenon [37], however, the impact of macroclimatic and local variables is evident in most of Colombia and many other regions in the world, as reported by WHO and the IPCC. Advances in understanding causality between climate, weather and dengue incidence can lead to an appropriate approach, both in time and in space, to design and implement preventive measures against potential dengue outbreaks and early warning systems based on ENSO and local climate variables. Also, our results contribute to understand and anticipate the possible consequences of climate change on this type of diseases.

We note that even if we considered the ’’natural regions” of Colombia to study the possible association between dengue cases, ENSO and climate variables, these regions were defined based on many variables, such as relief, climate, vegetation and soil type, that may not be determinant in the studied phenomenon. Besides, other relevant variables are not taken into account, such as urbanization, levels of immunity to the four serotypes of dengue related to past exposures, and age distribution. A more in-depth analysis should be made defining regions by grouping sites according to the most relevant variables in dengue dynamics, considering socioeconomic factors and the presence of other types of mosquitoes that compete with Ae. Aegypti.

## Conclusions

The relationship between climate, weather, human behavior, and arbovirus diseases is complex, making it difficult to identify the causal mechanisms. The determination of macro and microclimatic variables that influence dengue outbreaks and the temporal lags of their effects are extremely useful for the construction and implementation of early warning systems and preventive and control measures. Therefore, our results contribute to the development of climate-based surveillance, prevention and control programs for dengue fever in endemic areas of Colombia, shedding light on decision-makers about the adequate timing to implement prevention and control measures and to anticipate the effects of climate variability and climate change.

The spatial and temporal distribution of dengue fever cases in Colombia are associated with macroclimatic and local conditions. El Niño (La Niña) phenomenon is related to the intensification (weakening) of dengue cases at the national level and in most regions. Our results suggest that this association is mainly explained by ENSO-driven increase in temperature and decrease in rainfall. However, these associations are not simultaneous, and the temporal delay varies regionally.

The influence of ENSO and different climatic variables affected by this phenomenon vary in space and time, and is not stationary, so it is not easy to extrapolate results from one site to another. The association between dengue cases and ENSO varies when data are disaggregated by seasons. The Amazon region does not show a significant association when the complete data series is analyzed, although it appears at seasonal timescales, with high positive correlations in some of them. The opposite occurs in the Andes region, where the analysis of the complete time series indicates very high positive cross-correlations, which are reduced at seasonal timescales.

On the other hand, there are specific sites that control the relationship between dengue dynamics and ENSO at larger geographical/political spatial scales, e.g., what happens in the Pacific and Andes regions determine the relationship in Colombia, and the behavior of Antioquia and Valle del Cauca determine those of the Andes and Pacific regions, respectively.

The main difference between the linear (Pearson) and non-linear methods used in this work is that the former shows a strong cross-correlation between maximum temperature and the number of dengue cases (at the national level *ρ* =0.55, in the Andes region *ρ* =0.42, and in the Pacific region *ρ* =0.624), while the PCMCIplus method does not indicate a direct causality between these two variables (only an indirect causality in the Pacific region). This suggests that, if there is any effect of *T_max_* on the dengue dynamics on a national and regional scale, this is through other variables such as rainfall, relative humidity, and wind velocity. On the other hand, although the dynamic cross-correlogram (see 5) shows the time in which the association between ENSO and dengue cases starts to decrease, it does not quantify the degree of relationship for each period and time of the time-series, as does the wavelet transform.

Cross-correlation and wavelet analyses elude the real causalities involved in the climate-ENSO-dengue phenomenon, but the PCMCI method pursues this objective, confirming the effect of ENSO on precipitation and temperature, and their consequent effect on the number of dengue cases. However, none of these methods indicates the mechanisms underlying the association and relationships, so further studies are required to develop explanatory models and mechanistic analysis. Moreover, other macro-climate phenomena affecting the hydroclimatology of Colombia, such as the North Atlantic Oscillation, the Pacific Decadal Oscillation, the Atlantic Multidecadal Oscillation, the Madden-Julian Oscillation, among others [72], may alter the dengue dynamics, as well as socioeconomic factors and previous government interventions.

## Data Availability

We used data provided by the IDEAM - Instituto de Hidrología, Meteorología y Estudios Ambientales, and from the Instituto Nacional de Salud de Colombia.

https://www.ins.gov.co/buscador-eventos/Paginas/Info-Evento.aspx

http://geoservicios.ideam.gov.co/geonetwork/srv/spa/catalog.search#/home

## Supporting information

**S1 Fig.**
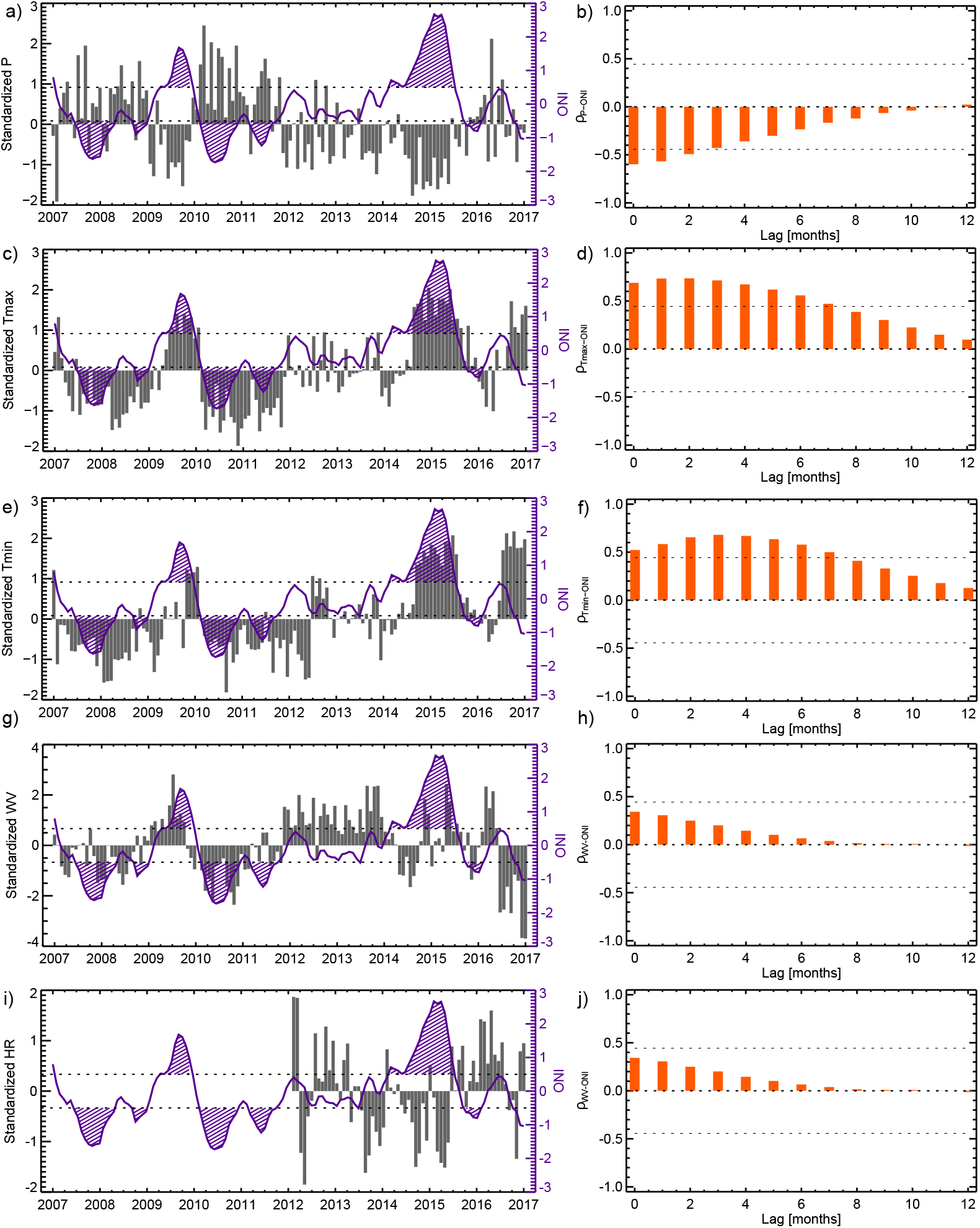
Time-series and cross-correlations of ONI and climate variables in Colombia. Time-series (left panel) and cross-correlations (right panel) between ONI and *P* (a-b), *T_max_* (c-d), *T_min_* (e-f), *WV* (g-h), and *HR* (i-j).

**S2 Fig.**
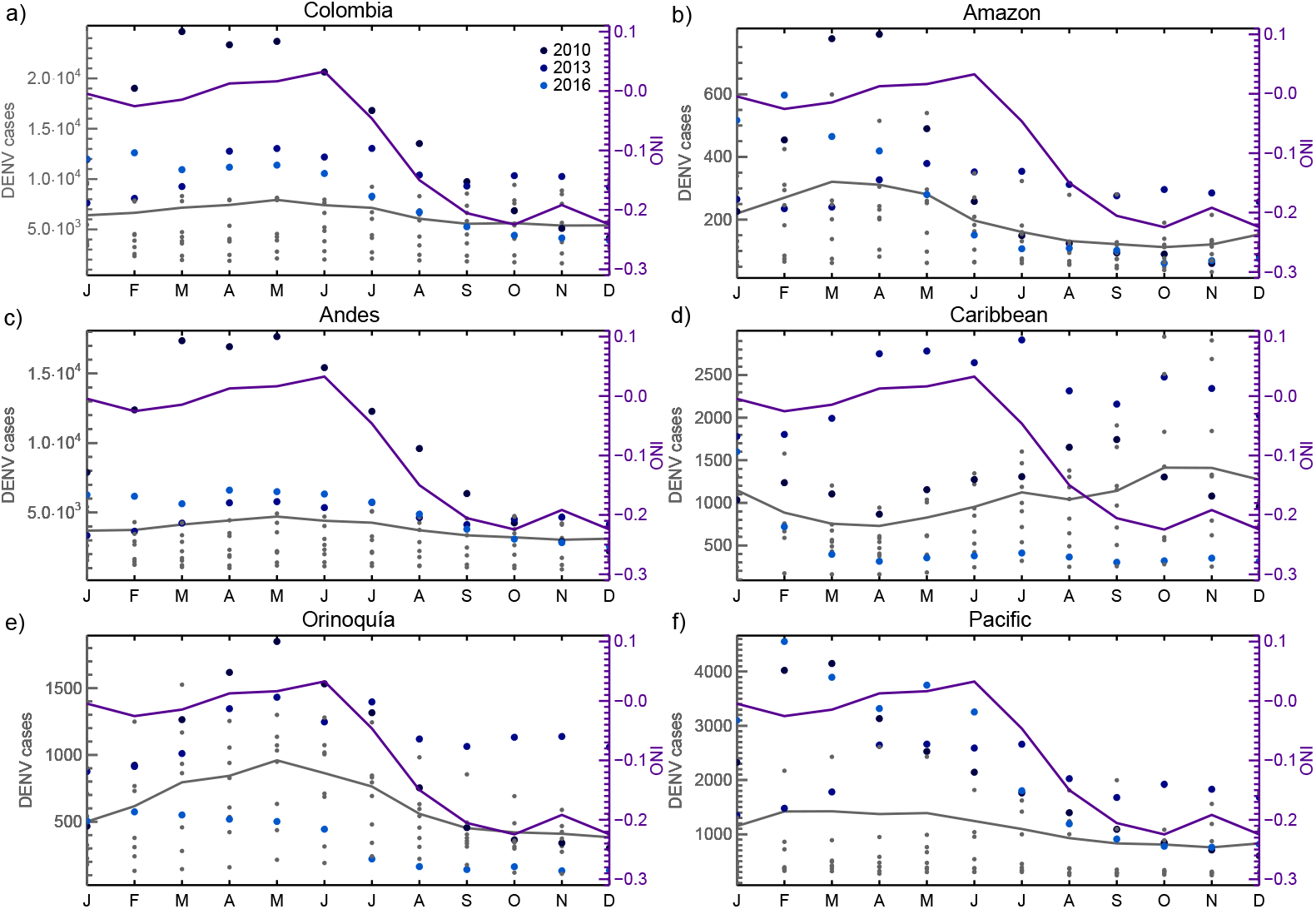
Monthly-annual cycles of ONI and dengue cases at the national and regional scales. Purple and gray solid lines indicate the annual-cycle of the ONI and dengue cases between 2007 and 2017, respectively. Blue dots denote the monthly number of cases in the hyperendemic years (shown in the legend), and the gray dots the number of cases in the other years.

## Acknowledgments

We are grateful to Mabel Calim Costa and Jakob Runge for making available the Waipy and Tigramite toolkits, respectively. We also thank the Instituto de Hidrología, Meteorología y Estudios Ambientales - IDEAM, and the Instituto Nacional de Salud de Colombia - INS, by the availability of the climatic and dengue data, respectively. The work of G. Poveda was supported by Universidad Nacional de Colombia at Medellín, Colombia, as a contribution to the project ”Detectión temprana de transmisión de dengue basada en diagnóstico molecular e informatión ambiental y climática en Santiago de Cali, Departamento Valle del Cauca, Colombia”, which was originally funded by COLCIENCIAS and currently by the Minister of Science, Technology and Innovation of Colombia. The work of E. Muøoz, M.P Arbeláez, and I.D. Vélez was supported by the World Mosquito Program, Colombia.

